# Modeling the Therapeutic Process of Patients with Cocaine Use Disorders: The Texas Christian University Process Model as a Guidance to Predict Readmission

**DOI:** 10.1101/2023.07.09.23292427

**Authors:** Daniel Dacosta-Sánchez, Fermín Fernández-Calderón, Carmen Díaz-Batanero, Cinta Mancheño-Velasco, Óscar M. Lozano

**Affiliations:** Department of Clinical and Experimental Psychology, University of Huelva, Huelva, Spain; Research Center on Natural Resources, Health and the Environment, University of Huelva, Huelva, Spain

**Author notes:** Correspondence to: Daniel Dacosta Sánchez. Department of Clinical and Experimental Psychology. University of Huelva. Facultad de Ciencias de la Educación. 21071 Huelva, Spain. EU.

**Keywords:** Therapeutic process, Texas Christian University Model, time in treatment, adherence of treatment, readmissions, successful/drop out treatment

## Abstract

**Background:** Patients readmitted for Cocaine Use Disorders are, along with Opiates Use Disorder, one of the group of patients with highest demand of treatment in specialized centers of addiction and with greater therapeutic failure.

**Objective:** Our aim is to apply the Texas Christian University Process Model to modeling the relationships between patient’s attributes at intake of treatment, treatment progress indicators and outcomes, including treatment success and readmissions.

**Method:** A retrospective observational design was used with 10,298 Cocaine Use Disorder patients. Electronic health records were used for statistical analysis of the data. Randomized subsample 1 (n= 5,150) was used for exploratory analysis and subsample 2 (n = 5,148) to modeling variables relations.

**Results:** Patients attributes at intake have limited relevance in explaining the treatment progress indicators and outcomes. Time on treatment and patient’s adherence are relevant to explain treatment success. Readmissions are mainly explained by time in treatment and therapeutic success. Been referred to addiction centers by health of services also appear to be relevant.

**Discussion and conclusion:** Our study reflects that the therapeutic process is important in order to have and adequate therapeutic adherence and to stay longer in treatment. Patients with a successful treatment and longer stay in treatment are less likely to have future readmissions. Through this study we highlight, therefore, the value of an adequate therapeutic adherence to obtain successful short- and medium-term results. This would make the treatment of these patients more efficient, and alleviate suffering for the patients and their families.

## Introduction

Substance use disorders (SUDs) are considered chronic disorders [1, 2], and it has been shown that their evolution resembles that of other chronic diseases such as hypertension, diabetes, and asthma [3]. Despite these similarities, from a therapeutic standpoint, SUDs are usually treated as acute disorders. While this approach can be useful in patients with non-severe diagnoses, for whom different interventions effectively reduce consumption and risk of relapse [4, 5], many SUD patients suffer repeated relapses that usually lead to treatment readmission, indicating that these treatments do not produce the expected effect. Reports published in Europe [6] and the United States [7] have revealed that more than 50% of treatment admissions correspond to patients who have previously undergone treatment, mainly patients with opioid and cocaine dependence. This phenomenon, also known as “revolving door” or “repeated treatment admission,” is associated with an increase in negative consequences for their physical health and quality of life [8], as well as an increase in healthcare costs [9].

Numerous studies have examined the factors associated with drug use relapse, either during treatment [10, 11] or after completion of treatment [12–14]. Relapse and subsequent hospital readmission resulting from severe drug-related consequences and detoxifications have also been analyzed [15–18]. In contrast, readmission to initiate new addiction treatment has been rather less studied. This type of readmission entails relapse in use and the patient’s own recognition of physical, psychological, and social deterioration, leading to the decision to seek therapeutic help again [4, 19]. Evidence from this line of research points to various factors that lead to readmission, which may differ according to the type of substance used. For example, studies using large samples of patients with substance use disorders concerning various drugs [20–22] point to certain patient characteristics more strongly associated with readmission, such as the type of substance used (e.g., amphetamine), being female, having been arrested, or having comorbid mental disorders. Studies conducted exclusively with opiate use disorder patients [23, 24] show that patients with greater post-treatment deterioration and certain sociodemographic characteristics and psychiatric comorbidities are more likely to require readmissions. Findings from authors who have studied patients with alcohol use disorder exclusively suggest that premature treatment dropout, psychiatric health problems, younger age, and longer duration of treatment are more strongly associated with readmission than other factors [25, 26]. In the case of patients with cocaine use disorder (CUD), Grella et al. [27] identified that using services in addiction centers, certain sociodemographic characteristics, and the pattern of use were all associated with readmission.

Taken together, the evidence indicates that readmissions are related to multiple factors that can be grouped into a) patient characteristics; b) human and material resources provided by addiction services; and c) other factors associated with the therapeutic process or activities [28–32].

These categories are captured within the Texas Christian University (TCU) Treatment Process Model [32], developed through decades of studying various addiction treatment programs. This model defines a set of sequential phases in a patient’s recovery and establishes how therapeutic interventions should be linked to these phases to maintain patient engagement and retention in the therapeutic program, which will result in patient improvement both during treatment and after its completion. Thus, this model highlights the importance of readiness for treatment and treatment engagement in achieving therapeutic success. In addition, the model states that patient variables at intake and program attributes can positively or negatively affect these variables. On the other hand, the model also points out the convenience of establishing observable indicators of treatment engagement, as these contribute to monitoring the therapeutic process and predicting outcomes [32, 33].

Since its definition, the TCU Treatment Process Model has been applied to various treatment programs, helping to identify the variables on which to focus interventions, while modifying organizational aspects to improve patient care and improve treatment effectiveness [33–35]. However, few studies have analyzed the relationships between the various indicators of the TCU Treatment Process Model with readmission to treatment. Moreover, considering the high heterogeneity among patients with SUD and the diversity of variables related to therapeutic programs, there is no consensus regarding which factors are determinant in explaining the effectiveness of the programs and the need for additional treatments. In particular, no studies have analyzed readmission to treatment under the TCU Treatment Process model in patients with cocaine dependence, in spite of the fact that these patients account for a high percentage of those admitted and readmitted to treatment each year in centers in the United States and Europe [6, 7].

Given this background, the present study had the following objectives: 1) to analyze which variables and indicators have the greatest explanatory power for each of the elements of the TCU Treatment Process Model in outpatients with cocaine use disorder; and 2) to evaluate the application of the TCU Treatment Process Model for explaining treatment readmission in outpatients with cocaine use disorder. Considering the evidence found in previous studies analyzing the variables associated with readmission, as well as the framework of relationships established by the TCU Treatment Process Model, it is hypothesized that: 1) a more severe pattern of use and the presence of psychiatric comorbidity will be negatively associated with patient engagement and retention in treatment [22, 23, 25]; 2) treatment outcomes will be influenced by how long patients are in treatment and by their engagement with treatment [32, 33]; 3) treatment outcomes will be the main explanatory factor for readmission to treatment [23, 24]; and 4) a longer time in treatment will be associated with a lower likelihood of readmission to treatment [25].

## Materials and Methods

### Design

This study adopted a retrospective observational design, with a follow-up period of 24 months from the start of treatment.

### Participants

The sample consisted of patients diagnosed with cocaine dependence according to ICD-10 criteria who began treatment in one of the 121 Public Network for Addiction Care centers in Andalusia (Spain) between 01/01/2015 and 12/31/2019. The Public Network for Addiction Care in Andalusia serves more than 95% of patients with addiction problems in this region (with more than 8 million inhabitants). This public network can be accessed at no financial cost to the patients, and each center is staffed by multidisciplinary teams of physicians, psychologists, nurses, and social workers. In addition, patients attending these centers follow cognitive-behavioral therapy [36].

To select the study sample, the following inclusion criteria were established: a) having been in treatment for at least one month; b) having at least one appointment for diagnostic assessment in addition to admission; c) not having been diagnosed with opiate abuse or dependence; d) not having been referred to another addiction resource; e) not having died while in treatment.

After applying these inclusion criteria, the sample consisted of 12,474 outpatients. Each patient was followed for 24 months. Therefore, the time frame of the study covered the period up to 31/12/2021. During the 24 months, 2,176 patients in treatment were excluded from the analyses. Thus, the final study sample consisted of patients who had received a therapeutic discharge (1,613 patients) and those who abandoned their treatment (8,685 patients), giving a total of 10,298 patients.

The study sample was 88.6% male. The mean age at the time of admission to treatment was 34.78 years (SD = 8.8), with no statistically significant differences between men and women. Of the patients, 55.7% had completed primary education, 40.3% had completed secondary education, while 4% had completed university studies. More than half (58.8%) of the patients were unemployed, 33.2% were working, 6.1% were pensioned, and the remaining percentage were studying. Regarding treatment, 67.3% had previously undergone treatment, and 48.5% had also been treated for cocaine use.

All patients were diagnosed with cocaine use disorder, while 47.3% had also been diagnosed with alcohol use disorder, and 31.1% with cannabis use disorder. In addition, 19.9% had a comorbid diagnosis of another mental disorder. Specifically, 11.1% had a personality disorder, 7.1% had an anxiety disorder, 3.8% had a mood disorder, 2% had attention deficit hyperactivity disorder (ADHD), and 2.4% had psychotic disorders.

Patients were in treatment for, on average, 9.9 (SD = 8.1) months. The mean number of scheduled appointments given to these patients during their treatment was 11.07 (SD = 11.88), and they showed an appointment attendance ratio of 0.43 (SD = 0.29) on average.

### Procedure

The data in the present study belong to the electronic health records (EHR) of the patients treated in public addiction centers in Andalusia. The EHR is registered through the Information System of the Andalusian Plan on Drugs (SiPASDA), which stores the information in a centralized database for all addiction centers. The EHR begins with recording information collected according to the standards set by the European Monitoring Centre for Drugs and Drug Addiction [37], including sociodemographic variables, drug use history, previous treatments, and infectious diseases. Then, this information is supplemented with clinical data during the patients’ routine appointments with clinical team members.

The EHR is automatically programmed to prevent missing data regarding important medical record variables. It can also detect errors and inconsistent response patterns, which contributes to the validity of the recorded data [38]. However, the information used in this study does not present missing values in any of the variables analyzed. This is because the program for collecting EHRs is designed so that clinicians are required to enter the relevant information on the study variables of the present work.

### Measures

According to the TCU Treatment Process Model, the following measures were used as patient attributes at intake:

- Sociodemographic data and variables related to the consumption patterns analyzed in this study correspond to those recorded in the treatment demand indicator (TDI) standard protocol 3.0 [39].
- Presence of comorbid mental disorders: The addiction centers’ clinicians made the diagnosis following ICD-10 diagnostic criteria.

Two indicators were calculated as a measure of patient engagement with treatment:

- Appointment attendance ratio: calculated by dividing the number of therapy sessions attended by the total number of sessions scheduled by the therapy team. A value of one indicates 100% attendance to scheduled appointments.
- Retention in treatment: calculated as the number of months from when the patient enters treatment until treatment ends.

The following measures were considered as indicators of treatment outcomes:

- Therapeutic outcome or short-term therapeutic outcome. Patients were classified according to whether they had achieved the therapeutic objectives (therapeutic success group) or whether they had dropped out of treatment (dropout group).
- Readmissions or long-term therapeutic outcome. Patients were considered readmissions when, after six months without attending the planned therapeutic appointments, the patient attended again, seeking the initiation of new treatment.

### Statistical Analysis

To address the study objectives, the global sample was divided into two subsamples, randomly selected using random numbers generated in SPSS vers. 24 [40]. First, to analyze which variables and indicators best explain each of the elements of the TCU Treatment Process Model, we used “subsample 1” (n=5,150), following an exploratory approach. Then, bivariate analyses were applied between the patient’s attributes at intake, treatment engagement indicators, and outcome variables. Given the large sample size, significant associations between variables were considered to be those that, in addition to showing a statistically significant association, had at least a “weak” effect size according to standard classifications [41]. Therefore, the effect size estimated by the Phi coefficient was used to analyze dichotomous variables, considering at least weak relationship from a value of φ ≤ .1 [41].

Second, to evaluate the application of the TCU Treatment Process Model in predicting readmission to treatment, “subsample 2” (n=5,148) was used using a confirmatory approach. For this purpose, a path analysis was applied, including as indicators in the model those variables with effect sizes at least low in the bivariate analysis conducted with subsample 1. Furthermore, to evaluate model fit, CFI values and TLI ≥ .90 are considered indicative of adequate fit; while RMSEA < 0.05 and SRMR < 0.08 are considered acceptable [42, 43]. Finally, to determine the contribution of the variables in the model, standardized regression coefficients, together with their confidence intervals, estimated by bootstrapping 1,000 samples, were evaluated. Statistical analyses were conducted with SPSS 24.0 [40] and MPlus 8.6 [44].

## Results

### The Contribution of Variables and Indicators to Explaining Progress and Treatment Outcomes

Tables 1 to 3 show the relationship between the appointment attendance ratio and patient attributes at intake. Among the sociodemographic variables (Table 1), patients with a university education attended a higher proportion of scheduled appointments. Patients who live with friends, as well as those who live with a family member with a history of addiction, attend fewer appointments, with an effect size greater than .20. Among the variables associated with the pattern of use (Table 2), patients with cannabis dependence attended a lower proportion of scheduled appointments (*d*= 0.245). Among the variables associated with dual pathology (Table 3), patients with mood disorders attended more appointments (Δ= 0.268).

**Table 1.** Proportion of appointments attended by patients according to sociodemographic and social characteristics

|  | Mean (SD) | *t* student | *d.f.* | *p* | Cohen's *d*/<br>Glass's delta |
| --- | --- | --- | --- | --- | --- |
| **Sex** |  |  |  |  |  |
| Male (n = 4459) | 0.44 (0.29) | 1.391 | 769.9 | .184 | Δ= 0.061 |
| Female (n=589) | 0.42 (0.27) |  |  |  |  |
| **Having children** |  |  |  |  |  |
| No (n=1974) | 0.44 (0.29) | 1.058 | 4040.5 | .290 | Δ= 0.031 |
| Yes (n=3174) | 0.43 (0.28) |  |  |  |  |
| **Highest educational level completed** |  |  |  |  |  |
| **Primary level** |  |  |  |  |  |
| No (n=2277) | 0.46 (0.29) | 5.231 | 5146 | .000 | *d*= 0.147 |
| Yes (n=2871) | 0.42 (0.29) |  |  |  |  |
| **Secondary level** |  |  |  |  |  |
| No (n=3079) | 0.42 (0.29) | 3.477 | 5146 | .000 | *d*= 0.099 |
| Yes (n=2069) | 0.45 (0.29) |  |  |  |  |
| **Higher education** |  |  |  |  |  |
| No (n=4940) | 0.43 (0.29) | 4.937 | 228.6 | .000 | *d*=0.320 |
| Yes (n=208) | 0.52 (0.26) |  |  |  |  |
| **Employment status** |  |  |  |  |  |
| **Unemployed** |  |  |  |  |  |
| No (n=2037) | 0.45 (0.30) | 3.457 | 4167.2 | .000 | Δ= 0.102 |
| Yes (n= 3111) | 0.42 (0.28) |  |  |  |  |
| **Employed** |  |  |  |  |  |
| No (n=3508) | 0.43 (0.28) | 2.636 | 3001.8 | .009 | Δ= 0.081 |
| Yes (n=1640) | 0.45 (0.30) |  |  |  |  |
| **Retired** |  |  |  |  |  |
| No (n=4838) | 0.43 (0.29) | 1.749 | 5146 | .800 | *d*= 0.102 |
| Yes (n=310) | 0.46 (0.28) |  |  |  |  |
| **Student** |  |  |  |  |  |
| No (n=5061) | 0.43 (0.29) | 0.260 | 5146 | .795 | *d*= 0.028 |
| Yes (n=87) | 0.44 (0.27) |  |  |  |  |
| **Living status** |  |  |  |  |  |
| **Alone** |  |  |  |  |  |
| No (n=4708) | 0.43 (0.29) | 0.750 | 5146 | .453 | *d*= 0.037 |
| Yes (n=440) | 0.44 (0.29) |  |  |  |  |
| **With partner/children** |  |  |  |  |  |
| No (n=3098) | 0.44 (0.29) | 1.646 | 5146 | .100 | *d*= 0.047 |
| Yes (n=2050) | 0.43 (0.29) |  |  |  |  |
| **With parents** |  |  |  |  |  |
| No (n=3004) | 0.43 (0.29) | 1.479 | 5146 | .139 | *d*= 0.042 |
| Yes (n=2144) | 0.44 (0.29) |  |  |  |  |
| **With friends** |  |  |  |  |  |
| No (n=5063) | 0.44 (0.29) | 2.366 | 5146 | .018 | *d*= 0.259 |
| Yes (n=85) | 0.36 (0.26) |  |  |  |  |
| **Living with family members with addiction** |  |  |  |  |  |
| No (n=4337) | 0.45 (0.29) | 6.198 | 5146 | .000 | $d = 0.230$ |
| Yes (n=811) | 0.38 (0.28) |  |  |  |  |
| *Source of referral* |  |  |  |  |  |
| Medical service |  |  |  |  |  |
| No (n= 4600) | 0.43 (0.29) | 4.102 | 5146 | .000 | $d = 0.185$ |
| Yes (n=548) | 0.48 80.30) |  |  |  |  |
| Social service |  |  |  |  |  |
| No (n=5058) | 0.43 (0.29) | 0.498 | 5146 | .618 | $d = 0.053$ |
| Yes (n=90) | 0.45 (0.28) |  |  |  |  |
| Legal Services |  |  |  |  |  |
| No (n=4898) | 0.43 (0.29) | 0.425 | 271.0 | .671 | $\Delta = 0.027$ |
| Yes (n=250) | 0.44 (0.31) |  |  |  |  |
| Referral from family or friends |  |  |  |  |  |
| No (n=4228) | 0.43 (0.29) | 0.697 | 5146 | .482 | $\Delta = 0.025$ |
| Yes (n=920) | 0.44 (0.29) |  |  |  |  |
| Self-referral |  |  |  |  |  |
| No (n=2273) | 0.46 (0.30) | 4.446 | 4783.6 | .000 | $\Delta = 0.128$ |
| Yes (n=2875) | 0.42 (0.28) |  |  |  |  |

**Table 2.** Proportion of appointments attended by patients according to drug use patterns and treatment history

|  | Mean (SD) | *t* student | *d.f.* | *p* | Cohen's *d*/<br>Glass's delta |
| --- | --- | --- | --- | --- | --- |
| Alcohol dependence |  |  |  |  |  |
| No (n=2727) | 0.44 (0.30) | 1.718 | 5130.4 | .086 | $\Delta = 0.050$ |
| Yes (n=2420) | 0.43 (0.28) |  |  |  |  |
| Cannabis dependence |  |  |  |  |  |
| No (n=3555) | 0.46 (0.29) | 8.120 | 5146 | .000 | $d = 0.245$ |
| Yes (n=1593) | 0.39 (0.28) |  |  |  |  |
| Tobacco dependence |  |  |  |  |  |
| No (n=4624) | 0.43 (0.29) | 0.227 | 5146 | .820 | $d = 0.010$ |
| Yes (n=524) | 0.44 (0.27) |  |  |  |  |
| Frequency of cocaine use 30 days before entering treatment |  |  |  |  |  |
| No (n=1163) | 0.44 (0.299) | 0.262 | 5146 | .793 | $d = 0.009$ |
| Yes (n=3985) | 0.43 (0.29) |  |  |  |  |
| Usual route of administration |  |  |  |  |  |
| Smoke/inhale (n=914) | 0.48 (0.28) | 5.246 | 1381.3 | .000 | $\Delta = 0.185$ |
| Sniff (n=4233) | 0.42 (0.29) |  |  |  |  |
| Ever previously treated |  |  |  |  |  |
| No (n=1685) | 0.43 (0.32) | 0.147 | 2936.7 | .883 | $\Delta = 0.005$ |
| Yes (n=3463) | 0.43 (0.27) |  |  |  |  |

**Table 3.** Proportion of appointments attended by patients according to dual pathology

|  | Mean (SD) | *t* student | *d.f.* | *p* | Cohen's *d*/<br>Glass's *delta* |
| --- | --- | --- | --- | --- | --- |
| Patients with a diagnosis of mood disorder |  |  |  |  |  |
| No (n=4928) | 0.43 (0.29) | 3.895 | 5146 | .000 | *d*= 0.268 |
| Yes (n=220) | 0.51 (0.30) |  |  |  |  |
| Patients with a diagnosis of anxiety disorder |  |  |  |  |  |
| No (n=4762) | 0.43 (0.29) | 2.071 | 462.3 | .039 | $\Delta$= 0.110 |
| Yes (n=386) | 0.46 (0.27) |  |  |  |  |
| Patients with a diagnosis of psychotic disorder |  |  |  |  |  |
| No (n=5029) | 0.43 (0.29) | 1.830 | 5146 | .067 | *d*= 0.170 |
| Yes (n=119) | 0.48 (0.29) |  |  |  |  |
| Patients with a diagnosis of personality disorder |  |  |  |  |  |
| No (n = 4565) | 0.43 (0.29) | 0.009 | 756.1 | .993 | $\Delta$= 0.000 |
| Yes (n = 583) | 0.43 (0.28) |  |  |  |  |
| Patients with a diagnosis of ADHD |  |  |  |  |  |
| No (n=5042) | 0.43 (0.29) | 0.327 | 112.1 | .744 | $\Delta$= 0.032 |
| Yes (n=106) | 0.43 (0.23) |  |  |  |  |

The relationships between retention and patient attributes at intake are shown in Tables 4 to 6. Among these variables, it is observed that those who live with friends spend less time in treatment, while those referred from legal services (court/probation/police) spend more time in treatment (Table 4). In addition, regarding consumption pattern variables (Table 5), patients with a previous treatment history spend less time in treatment (Δ= 0.340). Finally, analysis of comorbid mental disorders revealed statistically significant differences in time in treatment, although none of the patients showed a considerable effect size (Table 6).

**Table 4.** Time in treatment of patients according to sociodemographic and social characteristics

|  | Mean (SD) | *t* student | *d.f.* | *p* | Cohen's *d*/<br>Glass's delta |
| --- | --- | --- | --- | --- | --- |
| Sex |  |  |  |  |  |
| Male (n = 4459) | 8.67 (6.80) | 0.399 | 5146 | .690 | *d*= 0.017 |
| Female (n=589) | 8.79 (6.80) |  |  |  |  |
| Having children |  |  |  |  |  |
| No (n=1974) | 8.71 (6.78) | 0.253 | 5146 | .800 | *d*= .007 |
| Yes (n=3174) | 8.66 (6.81) |  |  |  |  |
| *Highest educational level completed* |  |  |  |  |  |
| Primary level |  |  |  |  |  |
| No (n=2277) | 8.60 (6.71) | 0.717 | 5146 | .474 | *d*= 0.020 |
| Yes (n=2871) | 8.74 (6.86) |  |  |  |  |
| Secondary level |  |  |  |  |  |
| No (n=3079) | 8.75 (6.87) | 0.902 | 5146 | .367 | *d*= 0.026 |
| Yes (n=2069) | 8.58 (6.89) |  |  |  |  |
| Higher education |  |  |  |  |  |
| No (n=4940) | 8.67 (6.79) | 0.441 | 5146 | .659 | *d*= 0.031 |
| Yes (n=208) | 8.88 (6.99) |  |  |  |  |
| *Employment status* |  |  |  |  |  |
| Unemployed |  |  |  |  |  |
| No (n=2037) | 9.28 (7.21) | 5.015 | 4026.7 | .000 | Δ= 0.153 |
| Yes (n= 3111) | 8.29 (6.49) |  |  |  |  |
| Employed |  |  |  |  |  |
| No (n=3508) | 8.37 (6.55) | 4.599 | 2930.9 | .000 | Δ= 0.134 |
| Yes (n=1640) | 9.34 (7.25) |  |  |  |  |
| Retired |  |  |  |  |  |
| No (n=4838) | 8.66 (6.78) | 0.931 | 5146 | .352 | *d*= 0.055 |
| Yes (n=310) | 9.03 (0.05) |  |  |  |  |
| Student |  |  |  |  |  |
| No (n=5061) | 8.68 (6.79) | 0.474 | 5146 | .636 | *d*= 0.051 |
| Yes (n=87) | 9.02 (7.01) |  |  |  |  |
| *Living status* |  |  |  |  |  |
| Alone |  |  |  |  |  |
| No (n=4708) | 8.61 (6.74) | 2.394 | 509.9 | .017 | Δ= 0.119 |
| Yes (n=440) | 9.48 (7.40) |  |  |  |  |
| With partner/children |  |  |  |  |  |
| No (n=3098) | 8.59 (6.70) | 1.187 | 5146 | .235 | *d*= 0.034 |
| Yes (n=2050) | 8.82 (6.93) |  |  |  |  |
| With parents |  |  |  |  |  |
| No (n=3004) | 8.77 (6.9) | 1.163 | 4720.6 | .245 | Δ= 0.033 |
| Yes (n=2144) | 8.55 (6.64) |  |  |  |  |
| With friends |  |  |  |  |  |
| No (n=5063) | 8.71 (6.80) | 2.736 | 5146 | .006 | *d*= 0.299 |
| Yes (n=85) | 6.68 (6.45) |  |  |  |  |
| With family members with addiction |  |  |  |  |  |
| No (n=4337) | 8.76 (6.84) | 1.938 | 5146 | .053 | d= 0.074 |
| Yes (n=811) | 8.26 (6.52) |  |  |  |  |
| *Source of referral* |  |  |  |  |  |
| Medical service |  |  |  |  |  |
| No (n= 4600) | 8.62 (6.74) | 1.719 | 665.4 | .086 | Δ= 0.077 |
| Yes (n=548) | 9.18 (7.23) |  |  |  |  |
| Social service |  |  |  |  |  |
| No (n=5058) | 8.67 (7.98) | 0.575 | 5146 | .566 | d= 0.061 |
| Yes (n=90) | 0.09 (6.69) |  |  |  |  |
| Legal Services |  |  |  |  |  |
| No (n=4898) | 8.58 (6.75) | 4.160 | 270.6 | .000 | Δ= 0.269 |
| Yes (n=250) | 10.57 (7.38) |  |  |  |  |
| Referral from family or friends |  |  |  |  |  |
| No (n=4228) | 8.64 (6.79) | 0.924 | 5146 | .355 | d= 0.034 |
| Yes (n=920) | 8.87 (6.84) |  |  |  |  |
| Self-referral |  |  |  |  |  |
| No (n=2273) | 9.13 (6.97) | 4.238 | 4760.8 | .000 | Δ= 0.122 |
| Yes (n=2875) | 8.32 (6.63) |  |  |  |  |

**Table 5.** Time in treatment of patients according to drug use patterns and treatment history

|  | Mean (SD) | *t* student | *d.f.* | *p* | Cohen's *d*/<br>Glass's delta |
| --- | --- | --- | --- | --- | --- |
| Alcohol dependence |  |  |  |  |  |
| No (n=2727) | 8.88 (6.85) | 2.219 | 5091.443 | .027 | $\Delta = 0.062$ |
| Yes (n=2420) | 8.46 (6.73) |  |  |  |  |
| Cannabis dependence |  |  |  |  |  |
| No (n=3555) | 8.94 (6.90) | 4.085 | 3291.422 | .000 | $\Delta = 0.125$ |
| Yes (n=1593) | 8.12 (6.53) |  |  |  |  |
| Tobacco dependence |  |  |  |  |  |
| No (n=4624) | 8.71 (6.82) | 1.043 | 5146 | .297 | $d = 0.048$ |
| Yes (n=524) | 8.39 (6.59) |  |  |  |  |
| Frequency of cocaine use 30 days before entering treatment |  |  |  |  |  |
| No (n=1163) | 8.94 (6.70) | 1.453 | 5146 | .146 | $d = 0.048$ |
| Yes (n=3985) | 8.61 86.82) |  |  |  |  |
| Usual route of administration |  |  |  |  |  |
| Smoke/inhale (n=914) | 8.69 (6.72) | 0.044 | 5146 | .965 | $d = 0.002$ |
| Sniff (n=4233) | 8.68 (6.81) |  |  |  |  |
| Ever previously treated |  |  |  |  |  |
| No (n=1685) | 9.97 (7.56) | 8.968 | 2853.001 | .000 | $\Delta = 0.304$ |
| Yes (n=3463) | 8.06 (6.30) |  |  |  |  |

**Table 6.**
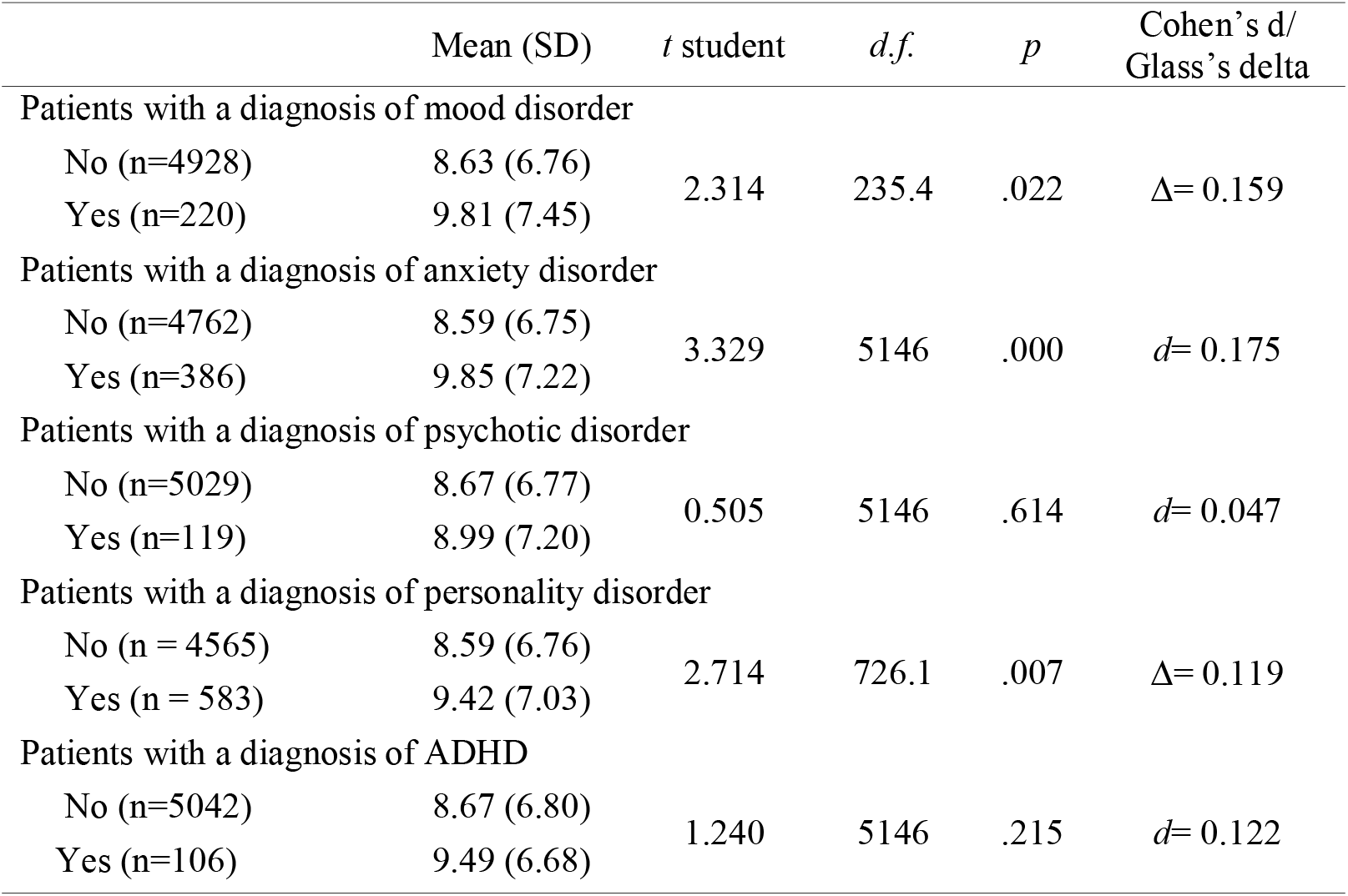
Time in treatment of patients according to dual pathology

|  | Mean (SD) | *t* student | *d.f.* | *p* | Cohen's *d*/<br>Glass's *delta* |
| --- | --- | --- | --- | --- | --- |
| Patients with a diagnosis of mood disorder |  |  |  |  |  |
| No (n=4928) | 8.63 (6.76) | 2.314 | 235.4 | .022 | Δ= 0.159 |
| Yes (n=220) | 9.81 (7.45) |  |  |  |  |
| Patients with a diagnosis of anxiety disorder |  |  |  |  |  |
| No (n=4762) | 8.59 (6.75) | 3.329 | 5146 | .000 | *d*= 0.175 |
| Yes (n=386) | 9.85 (7.22) |  |  |  |  |
| Patients with a diagnosis of psychotic disorder |  |  |  |  |  |
| No (n=5029) | 8.67 (6.77) | 0.505 | 5146 | .614 | *d*= 0.047 |
| Yes (n=119) | 8.99 (7.20) |  |  |  |  |
| Patients with a diagnosis of personality disorder |  |  |  |  |  |
| No (n = 4565) | 8.59 (6.76) | 2.714 | 726.1 | .007 | Δ= 0.119 |
| Yes (n = 583) | 9.42 (7.03) |  |  |  |  |
| Patients with a diagnosis of ADHD |  |  |  |  |  |
| No (n=5042) | 8.67 (6.80) | 1.240 | 5146 | .215 | *d*= 0.122 |
| Yes (n=106) | 9.49 (6.68) |  |  |  |  |

Regarding short-term outcomes, the results indicate that 84.4% of patients had prematurely dropped out of treatment, and 15.6% reached their therapeutic goals. None of the patient attributes at intake showed a considerable effect size regarding the association with therapeutic discharge/dropout. However, the treatment engagement variables showed a relationship with therapeutic discharge/dropout, with high effect sizes for time in treatment and appointment attendance ratio (Δ= 1.325 and Δ= 0.523, respectively).

Regarding readmission, after 24 months from the start of treatment, 27.4% of the patients were readmitted for treatment. Patients who had dropped out presented more readmissions than those who had not received a therapeutic discharge (29.9% vs. 13.9%; χ^2^ = 87.070; φ = 0.130). It is also observed that patients with high relapse rates spend less time in treatment with a high effect size (x = 9.95 (SD = 7.32) vs. x = 5.33 (SD = 0.36); p =.000; Δ= 1.374), and also attend a significantly lower proportion of appointments (x = 0.44 (SD = 0.30) vs. x = 0.42 (SD = 0.26); p =.002), although the effect size was not considerable (Δ= 0.101).

Among the patient attributes at intake, those who are employed, those initially referred to treatment by medical services, and those with a previous history of treatment showed fewer relapses (φ < 0.1).

### TCU Treatment Process Model Application for Predicting Readmission

Table 7 displays the results of applying the TCU Treatment Process Model, including the variables with an effect size greater than Cohen’s d/Glass’s delta higher than .20 in the previous exploratory analysis (Model 1). This model showed an acceptable fit (CFI = .960; TLI = .934; RMSEA = .027; RMSEA 90%CI = .022 - .032; SRMR = .028). However, analysis of the coefficients revealed that the variable “living with friends” yielded coefficients that were not statistically significant. This variable was excluded from a new model for the purposes of parsimony (Model 2). Model 2 showed similar fit indicators (CFI = .962; TLI = .939; RMSEA = .028; RMSEA 90% CI = .022 - .033; SRMR = .029).

**Table 7.** Comparison of fit statistics applying the TCU Treatment Process Model

| Variables | Model 1 |  |  | Model 2 |  |  |
| --- | --- | --- | --- | --- | --- | --- |
|  | Estimate | S.E. | p-value | Estimate (Int. Conf.)* | S.E. | P-value |
| *Readmissions 24 months: $R^2 = 0.394$, $p = .000$* |  |  |  |  |  |  |
| Drop out /Therapeutic success | 0.112 | 0.031 | .000 | 0.113 (0.037 – 0.185) | 0.031 | .000 |
| Retention in treatment | -0.419 | 0.029 | .000 | -0.421 (-0.487 - -0.362) | 0.025 | .000 |
| Medical service | -0.121 | 0.023 | .000 | -0.120 (-0.185 - -0.066) | 0.023 | .000 |
| Ever previously treated | 0.642 | 0.021 | .000 | 0.642 (0.592 – 0.699) | 0.021 | .000 |
| *Drop out /Therapeutic success: $R^2 = .673$, $p = .000$* |  |  |  |  |  |  |
| Time in treatment | 0.565 | 0.016 | .000 | 0.565 (0.521 – 0.602) | 0.016 | .000 |
| Proportion of appointments | 0.194 | 0.018 | .000 | 0.194 (0.146 – 0.240) | 0.018 | .000 |
| *Appointment attendance ratio: $R^2 = .017$, $p = .000$* |  |  |  |  |  |  |
| Higher education | 0.038 | 0.015 | .010 | 0.038 (0.003 – 0.073) | 0.015 | .010 |
| Living with friends | -0.003 | 0.150 | .738 | - | - | - |
| Living with family members with addiction | -0.073 | 0.014 | .000 | -0.073 (-0.108 - -0.037) | 0.014 | .000 |
| Cannabis dependence | -0.080 | 0.014 | .000 | -0.080 (-0.118 - -0.08) | 0.014 | .000 |
| Patients with a diagnosis of mood disorder | 0.052 | 0.014 | .000 | 0.052 (0.014 - 0.084) | 0.014 | .000 |
| *Time in treatment: $R^2 = .00439$, $p = .000$* |  |  |  |  |  |  |
| Living with friends | -0.010 | 0.016 | .521 | - | - | - |
| Legal services | 0.110 | 0.014 | .000 | 0.111 (0.059 – 0.149) | 0.014 | .000 |
| Ever previously treated | -0.156 | 0.014 | .000 | -0.157 (-0.201 - -0.116) | 0.014 | .000 |
| Fit Statistics | CFI = .960 |  |  | CFI = .962 |  |  |
|  | TLI = .934 |  |  | TLI = .939 |  |  |
|  | RMSEA = .027 (CI = .022 - .032) |  |  | RMSEA = .028 (CI = .022 - .033) |  |  |
|  | SRMR = .028 |  |  | SRMR = .029 |  |  |

*Note.* \*: Confidence interval estimated from a Bootstrap simulating 1,000 samples.

Among the patient attributes at intake, concerning time in treatment, referral by “legal services” predicts a longer time in treatment, while having previous treatment history predicts a shorter time in treatment. Respect to appointment attendance, cannabis dependence and living with family members with addictions were negatively associated with the appointment attendance ratio, while having a university education and suffering from mood disorders predicted a higher rate of treatment attendance (Figure 1). Regarding the indicators of therapeutic progress, both the appointment attendance ratio and time in treatment predict treatment outcomes (dropout vs. therapeutic discharge). Finally, the model shows that spending more time in treatment and achieving therapeutic objectives are predictors for not requiring readmission during the subsequent 24 months, while those with a previous treatment history are more likely to seek treatment again. Finally, of the patients who relapse, those referred by medical health services are less likely to return to treatment.

**Fig. 1.**
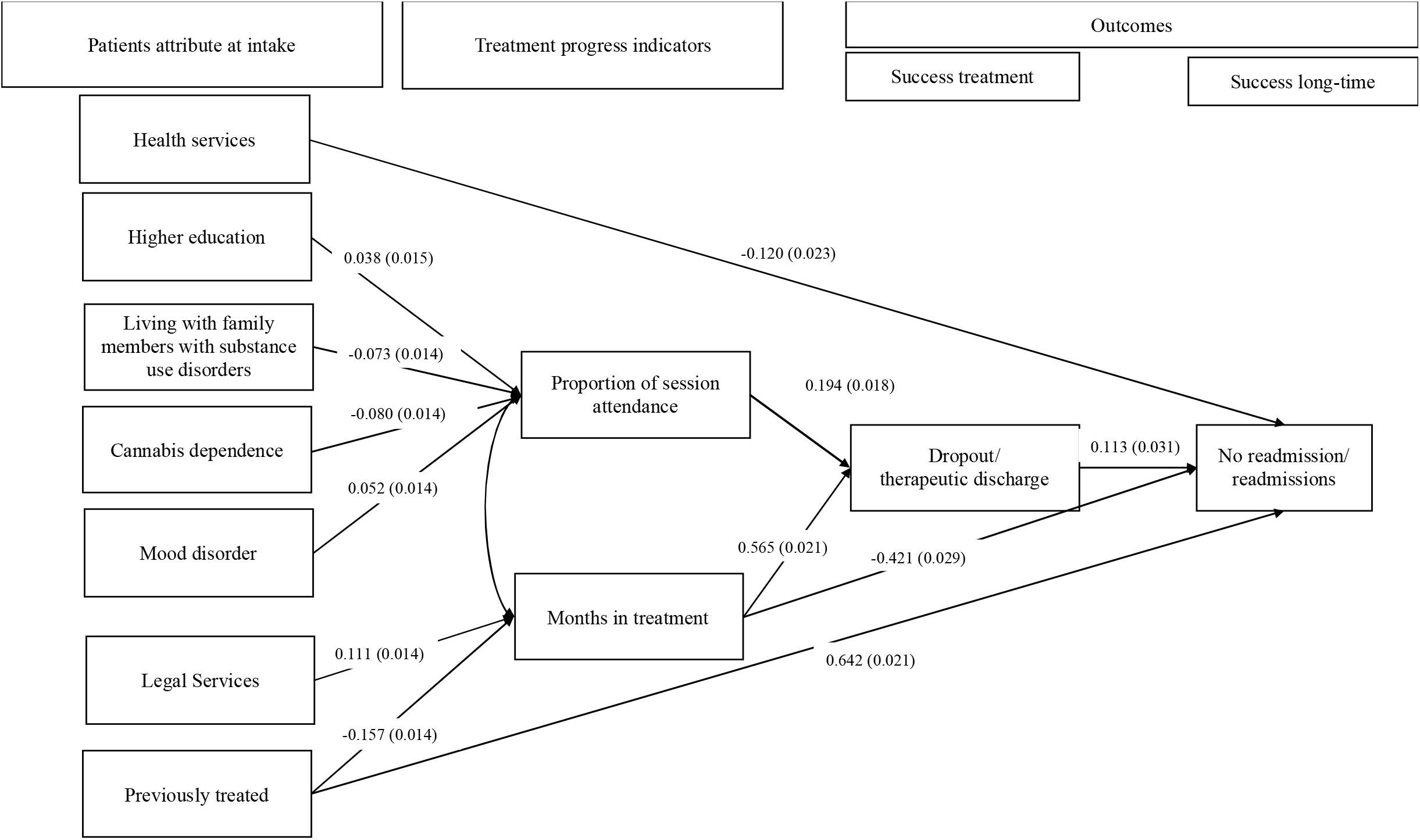
Path analysis applying the TCU Treatment Process Model

## Discussion and Conclusion

This study aimed to provide new evidence on how the TCU Treatment Process contributes to a more comprehensive framework for treating patients readmitted with cocaine dependence. This group of patients is currently one of the most demanding, largely due to readmissions [6, 7].

The literature has revealed mixed results when exploring the relationships between different variables (e.g., sociodemographic, consumption, dual pathology, therapeutic progress) and treatment outcomes [45–47]. In addition, the existing results are sometimes contradictory in the sense that the same variable can predict treatment outcomes in one study [48] and have no capacity for predicting outcomes in another [49], questioning the replicability of these results. These findings are difficult to explain, and it is likely that the differences between studies can explained the observed heterogeneity. Therefore, in this study, and considering the sample size, the analytical strategy was divided into two phases, following the recommendations of some authors [50]. A first exploratory phase was used to determine the relevant relationships between variables, using the effect size in those variables that were statistically significant; and in a second confirmatory phase, the significant relationships were modeled according to the TCU Treatment Process Model proposal.

The first aspect to highlight is that, generally, the baseline indicators associated with patient attributes are not predictive of treatment outcomes, according to the fixed effect size criterion [41]. However, patient attributes have predictive capacity for therapeutic progress indicators (months in treatment and proportion of scheduled appointments attended). This finding could be taken to indicate two possibilities. First, current treatments could be adapted to the personal characteristics of each patient so that these do not determine therapeutic outcomes and are instead modifiable by the treatments. Second, and in congruence with the TCU Treatment Process model and as shown by other authors [19, 51], time in treatment is a determinant of therapeutic success [32,52], while the attendance appointment ratio has also emerged as a relevant variable for assessing therapeutic success. In this sense, both variables have explained a high percentage of variance in the type of discharge (dropout vs. therapeutic success) in a period of 24 months. However, in congruence with the TCU Treatment Process Model, the coefficients show that time in treatment is a more significant predictor than the appointment attendance ratio. Thus, although both variables are useful indicators of the therapeutic process for the prognosis of patient outcomes, the time patients are in treatment appears to be the most relevant. Moreover, this variable and treatment outcome predict future readmission, as observed in alcohol-dependent patients [25].

Two other variables that have been shown to predict readmission are having a previous history of treatment and being referred to addiction treatment by medical services. Regarding the former variable, our study has shown that this is associated with the time in treatment (patients with previous admissions spend less time in treatment), and other studies have shown its relationship with treatment outcomes [53, 54]. Regarding the latter variable, the observation that readmission is less common among those who have been referred by a clinician could reflect the fact that healthcare professionals have greater authority or influence than others in promoting the benefits of starting addiction treatment.

Although the present study highlights variables that are of interest to clinicians and researchers for improving patient treatments, we believe it is necessary to highlight some limitations. Possibly the most important limitation concerns the study design. Real-world data have the advantage of providing information obtained by retrospective observation of a large volume participants. However, this type of design relies primarily on statistical analysis due to the sample size and indicates what has occurred over a period of time. And while the reliability and validity of the data in this study have been largely confirmed [55], a major flaw of this type of study is that there is little control over the research design [22]. In this regard, findings from this type of approach should be considered complementary to the evidence obtained through other research methods (and never as a substitute). Moreover, the gender distribution of the sample is highly asymmetrical, which casts doubts over the generalizability of these findings to women. This is a common limitation in most addiction research conducted with patients since there is a gender imbalance in treatment centers, with fewer women than men [37, 49]. However, given the sample size, other studies conducted by this research group focusing on gender differences have not been provided because we consider this to be beyond the scope of this study.

On the other hand, there may be a statistical effect (dropouts spend less time in treatment and, therefore, have more time for relapse), and the results obtained in this study are congruent with those found in the readmission of patients with alcohol dependence [25].

## Data Availability

All data produced in the present study are available upon reasonable request to the authors

## Acknowledgement

This study has been developed thanks to the transfer of data by the Department of Equality, Social Policies and Conciliation of the Junta de Andalucía.

## Statement of Ethics and Approvals

The storage and encoding of this data comply with the General Health Law of April 25, 1986 (Spain) and Law 41/2002 of November 14 on patient autonomy, rights, and obligations regarding clinical information and documentation. Furthermore, according to European regulations, the procedures comply with the Organic Law 3/2018 of December 5, 2018, on protecting personal data and guaranteeing digital rights. The Research Ethics Committee of the Andalusian Ministry of Health certified compliance with the ethical handling of the information.

## Author Contributions

Daniel Dacosta-Sánchez, Fermin Fernández-Calderón and Óscar M. Lozano have been implicated in the conceptualization, methodology, and study design. Daniel Dacosta-Sánchez, Carmen Díaz-Batanero, Cinta Mancheño-Velasco and Óscar M. Lozano have been involved in data curation and formal analysis.

All authors contributed to the drafting and revision of the manuscript.

